# Intra-operative Laxity Following Total Knee Arthroplasty is Highly Variable and Different Than Osteoarthritic and Normal Knees

**DOI:** 10.1101/2020.10.13.20212159

**Authors:** Robert A. Siston, Erin E. Hutter, Joseph A. Ewing, Rachel K. Hall, Jeffrey F. Granger, Matthew D. Beal

## Abstract

**Background:** Achieving a stable joint is an important yet challenging part of total knee arthroplasty (TKA). Neither manual manipulation of the knee nor instrumented sensors biomechanically characterize knee laxity or objectively characterize how TKA changes the laxity of an osteoarthritic (OA) knee. Therefore, the purposes of this study were: 1) objectively characterize changes in knee laxity due to TKA, 2) objectively determine whether TKA resulted in equal amounts of varus-valgus motion under a given load (i.e., balance) and 3) determine how TKA knee laxity and balance differ from values seen in non-osteoarthritic knees.

**Methods:** Two surgeons used a custom navigation system and intra-operative device to record varus-valgus motion under quantified loads in a cohort of 31 patients (34 knees) undergoing primary TKA. Similar data previously were collected from a cohort of 42 native cadaveric knees.

**Results:** Performing a TKA resulted in a “looser knee” on average, but great variability existed within and between surgeons. Under the maximum applied moment, 20 knees were “looser” in the varus-valgus direction, while 14 were “tighter”. Surgeon 1 generally “loosened” knees (OA laxity 6.1°±2.3°, TKA laxity 10.1°±3.6°), while Surgeon 2 did not substantially alter knee laxity (OA laxity 8.2°±2.4°, TKA laxity 7.5°±3.3°). TKA resulted in balanced knees, and, while several differences in laxity were observed between OA, TKA, and cadaveric knees, balance was only different under the maximum load between OA and cadaveric knees.

**Conclusions:** Large variability exists within and between surgeons suggests in what is considered acceptable laxity and balance of the TKA knee when it is assessed by only manual manipulation of the leg. Knees were “balanced” yet displayed different amounts of motion under applied load.

**Clinical Relevance:** Our results suggest that current assessments of knee laxity may leave different patients with biomechanically different knees. Objective intra-operative measurements should inform surgical technique to ensure consistency across different patients.

**Level of Evidence:** Level II prospective observational study

## Introduction

The success of total knee arthroplasty (TKA) depends on many factors, but surgical technique, including establishing a stable joint, has been identified as particularly critical.[1, 2] Improper intra-operative management of soft tissues can lead to post-operative complications such as instability, early loosening of the components, stiffness, limited range of motion, and excessive polyethylene wear.[3, 4] However, debate exists regarding how much joint laxity is appropriate. Some surgeons aim for a post-operative knee that is not too tight with a little varus-valgus laxity, looser in flexion than in extension, and looser laterally (under varus stress) than medially.[5-7] Similarly, patients have reported they prefer a lax knee compared to an over-tight knee.[8] Even though healthy knees are reported to have asymmetric mediolateral laxity[9], many surgical protocols advocate knees with equal amounts of varus-valgus laxity (i.e., a balanced knee). Ultimately, an objective definition of what constitutes a clinically successful amount of joint laxity does not exist.

Multiple techniques are used to assess joint laxity during TKA, but each has notable shortcomings. Many surgeons have become skilled at developing a “feel” for acceptable knee laxity during manual manipulation of the leg, but this qualitative evaluation lacks objective measurements and has unknown repeatability. Navigation systems have been used to measure varus-valgus range-of-motion and measure tibiofemoral spacing at the time of surgery[10-16], but the clinical benefit of navigation remains heavily debated. [11-14, 17, 18] Instrumented sensors record tibiofemoral contact to determine the balance provided by the collateral ligaments, with some studies suggesting that balanced intra-operative readings lead to improved clinical outcomes[19, 20] and post-operative kinematics [21]. However, these tibiofemoral contact readings are typically only recorded at one varus-valgus angle, and a variety of often conflicting tibiofemoral force targets have been reported. [19, 20, 22-24]

No current intra-operative approach to measuring knee laxity provides its historically proposed biomechanical characterization, specifically “continuous recordings of force-displacement and moment-rotation relationships”[25]. Previous studies have characterized knee laxity this way in healthy subjects [26, 27], in intact cadaveric specimens[25, 28], and in cadaveric specimens after TKA [29]. However, such objective measurements of TKA knee laxity have not been conducted intraoperatively and could help determine what magnitudes of knee laxity and balance best lead to improved post-operative function.

We believe the ability to objectively characterize intra-operative knee laxity would represent an improvement over the subjective or incomplete measurements currently made with existing approaches. We previously created a novel device that interfaces with our custom surgical navigation system to allow surgeons to quantify knee laxity [30]. In this study, we used those custom tools to 1) objectively characterize changes in knee laxity due to TKA, 2) objectively determine whether TKA resulted in equal amounts of varus-valgus motion under a given load (i.e., balance) and 3) determine how TKA knee laxity and balance differ from values seen in non-osteoarthritic knees. As a secondary analysis, we investigated these intra-operative data between two orthopaedic surgeons to assess the variability within and between surgeons.

## Methods

This study followed a protocol that was approved by the Institutional Review Board at The Ohio State University, and all participants provided written informed consent. Increased surgical time associated with our intra-operative measurements was limited to 20 minutes, and the surgeons (Beal and Granger) could elect to stop testing at any time. In this manuscript, we are reporting on a cohort of 31 participating patients (11 males, 20 females, average age 60.4±8.4 years) including 3 bilateral patients (all women) for a total of 34 knees (Beal: n=15, Granger: n=19), all with a pre-operative varus deformity.

We used an intra-operative measurement system comprised of a custom surgical navigation and device to objectively measure knee laxity [30-32]. After exposing the knee with a standard medial parapatellar arthrotomy, the surgeons placed threaded couplers in the tibia and femur to rigidly hold an optical tracker in each bone and then established reference frames by digitizing appropriate landmarks [33]. The laxity device was then placed on the table above the sterile drapes, the modified boot was applied to the patient’s foot using Coban wrap (3M, St. Paul, MN), and the boot was placed in the slider mechanism with the leg in full extension (Figure 1). To record laxity data, the surgeon placed the instrumented handle in the slider (Figure 1B) and restrained the femur by gripping the patient’s distal thigh with the knee in full extension. Without pre-conditioning the knee, the surgeon then applied a varus-valgus load for 3 trials as the navigation system recorded the resultant motion of the tibia, femur, boot, and slider and the instrumented handle measured the input load applied by the surgeon.

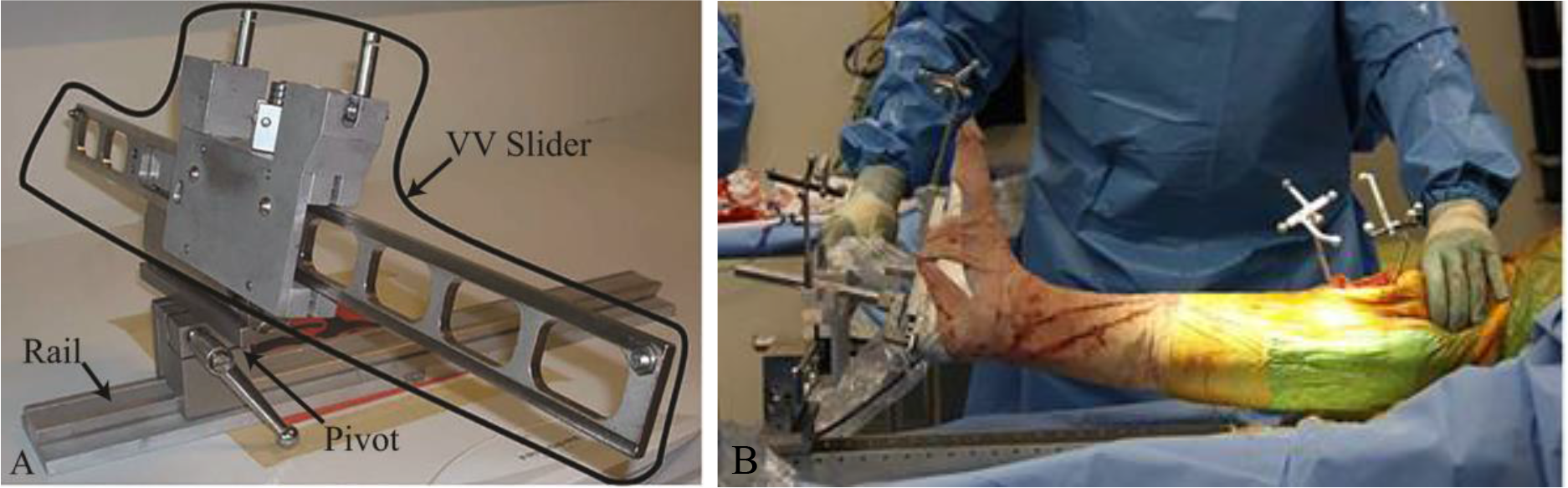
The custom laxity device has 5 main parts: varus-valgus (VV) slider, pivot, and rail, as seen in Figure 1A. Used in conjunction with the instrumented handle, the surgeon can collect varus-valgus laxity data in the operating room when the foot is placed in a modified Alvarado boot, as seen in Fig. 1B

The surgeons were blinded to the intra-operative measurements and were entirely in control of the amount of force applied to the patient. This approach, as apposed to prescribing a load that needed to be achieved for all patients, more closely resembled current clinical practice and allowed us to determine the range of laxity that was considered acceptable by our surgeons. Maximum loads varied based upon what the surgeon felt was safe for each patient; the maximum varus moment ranged from 0.3-35.2 N·m, and the maximum valgus moment ranged from 3.6-34.9 N·m.

Following testing on the osteoarthritic knee, surgeons then performed a TKA with a Zimmer NexGen LPS flex knee (Warsaw, IN) with similar surigical techniques. They both sought to approximate a neutral mechanical axis of the leg by making a valgus cut to the distal femur and a perpendicular cut to the proximal tibia. In the sagittal plane, cuts were designed to match the J-curve of the femur and establish a posterior slope on the tibia. The femoral component was rotationally aligned parallel to the transeipcondylar axis, while the tibial component was aligned to the medial third of the tubercle. Minimal soft tissue release was performed following bone cuts, although release of the deep medial collateral ligament off of the edge of the tibia is part of the initial surgical exposure. After the implants were cemented in the knee, the patient’s leg was put in the device, and varus-valgus data were taken again using the same procedure as before.

Intra-operative data were used to characterize knee laxity by plotting the moment applied to the leg versus the displacement (in degrees) for each pre- and post-implant trial (Figure 2). A third-order polynomial was fit to the raw data for each trial of alternating varus/valgus load. Each patient’s overall varus/valgus laxity was determined from the average of these three trials (Figure 3). The neutral position of the leg was established to be the varus/valgus angle of the knee under no load, even if this this angle was not 0°. We defined varus/valgus laxity as the amount of varus/valgus displacement from the neutral position that occurs under a given varus/valgus load. While applied moments of different values have previously been used to assess knee laxity [34-37], we analyzed laxity data at 2 different levels: 1) a 7.5 N·m varus/valgus moment [37, 38] (±7.5N·m laxity) and 2) the maximum varus/valgus moment applied to the limb (max varus/valgus laxity). We also investigated laxity in only one direction under both the 7.5 N·m and maximum applied loads (e.g., varus 7.5 N·m laxity) For the two knees that did not experience at least one trial of a **±**7.5 N·m moment, only their maximum moment was calculated.

**Figure 2:**
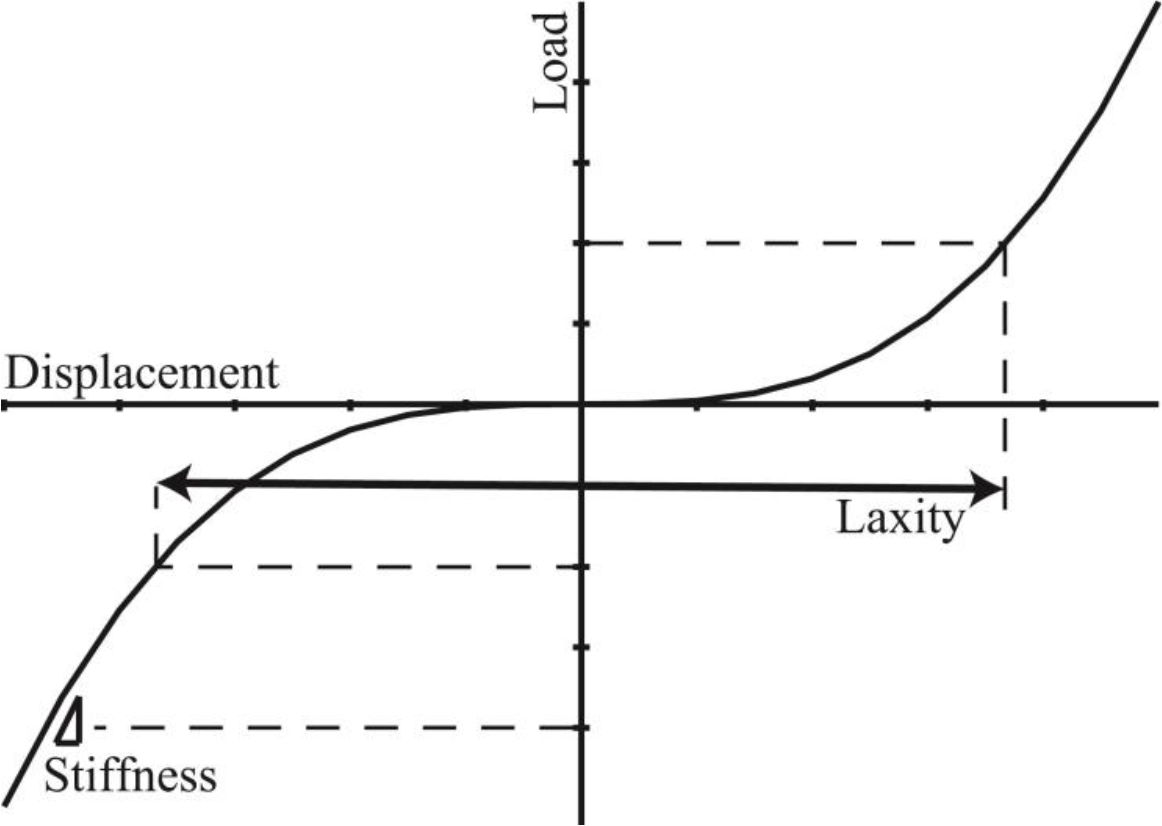
Load-Displacement curves were used to analyze the intra-operative data. Laxity is the displacement under a given load.

**Figure 3:**
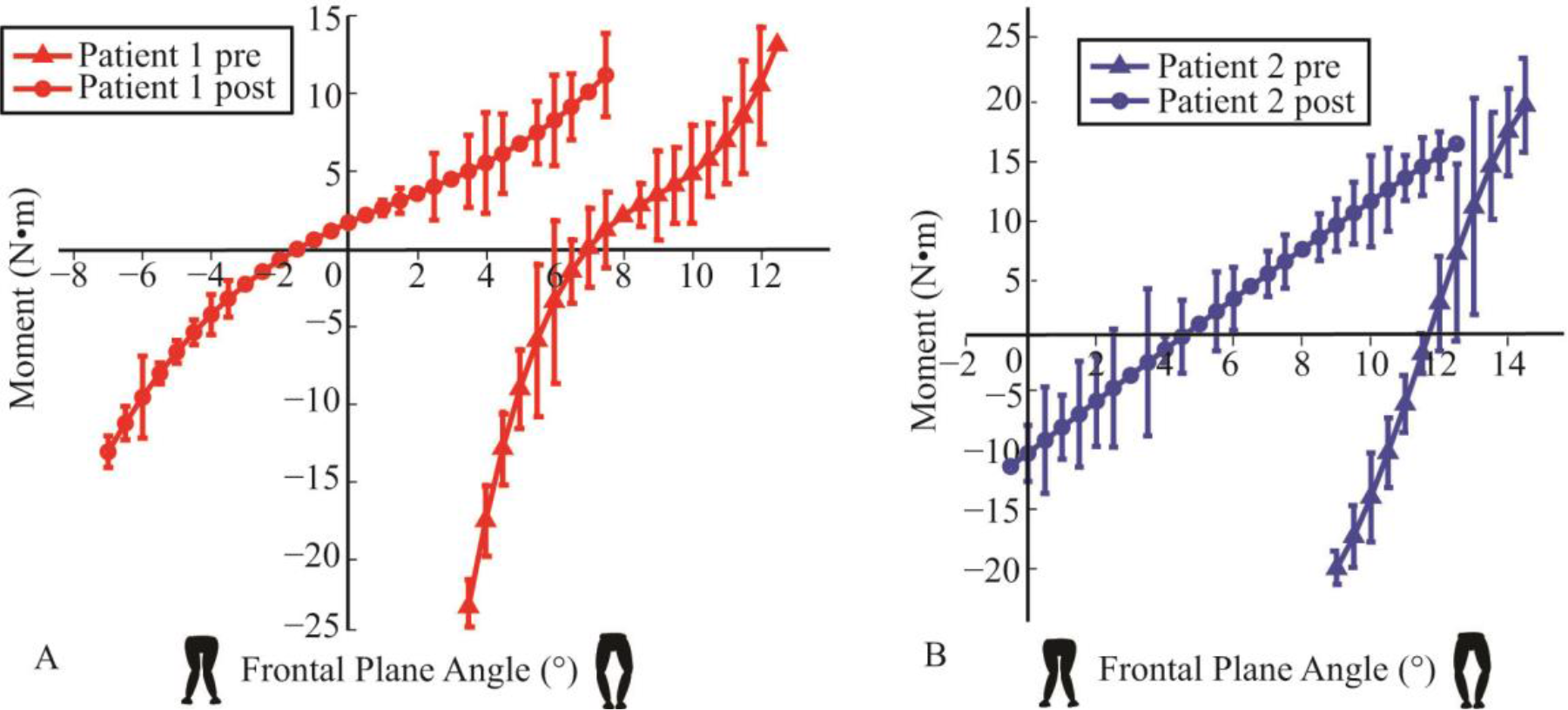
Representative varus-valgus laxity data from two representative patients (1 and 2) with the knee in full extension. Mean values from three cycles of testing are plotted along with ± one standard deviation.

A dataset of “normal” joint laxity was estaslished from similar measurements in 42 cadaveric specimens that were part of ongoing research in our laboratory (20 from female donors, average age 69.0 ± 14.9 years, age range: 27-87 years). These cadaveric knees had observed levels of OA of “none” or “mild” as determined by participating orthopaedic surgeons. While cadavers cannot be considered a perfect representation of “normal” anatomy, the need to drill trackers into the bones with the intra-operative device renders it inappropriate for use in healthy volunteers.

We employed several statistical analyses. In order to characterize the change in knee laxity from the pre-operative osteoarthritic knee to the knee after TKA and to determine whether the individual surgeons affected post-operative laxity, a general linear model ANOVA was used to test for significance of surgeon, OA/TKA knee condition, and the surgeon*OA/TKA interaction effect. Upon seeing a significant interection effect in the ANOVA, we analyzed magnitudes of OA/TKA laxity on each patient for each surgeon using the paired t-test. One-sample t-tests were used to test for differences in mediolateral balance in the postoperative (TKA) knees by comparing the difference in varus and valgus laxity to “0”, which would suggest a “perfectly balanced” knee with equal amounts of varus/valgus laxity. Two sample t-tests were used to test for differences in the magnitudes of laxity and balance between surgeons. Separate two sample t-tests with Bonferroni corrections were used to compare both intraoperative OA and TKA knee laxity data with corresponding data from the cohort of native cadaveric knees.

## Results

### Objectively characterize changes in knee laxity due to TKA

Performing a TKA resulted, on average, in a “looser” knee in the varus-valgus direction, (Tables 1-4). However, there was great variability in the results. Seventeen knees were “looser” in the varus-valgus direction after TKA compared to before TKA, while 15 were “tighter” with a ±7.5 N·m applied load. Under the maximum applied moment, twenty knees were “looser”, while 14 were “tighter”.

**Table 1:**
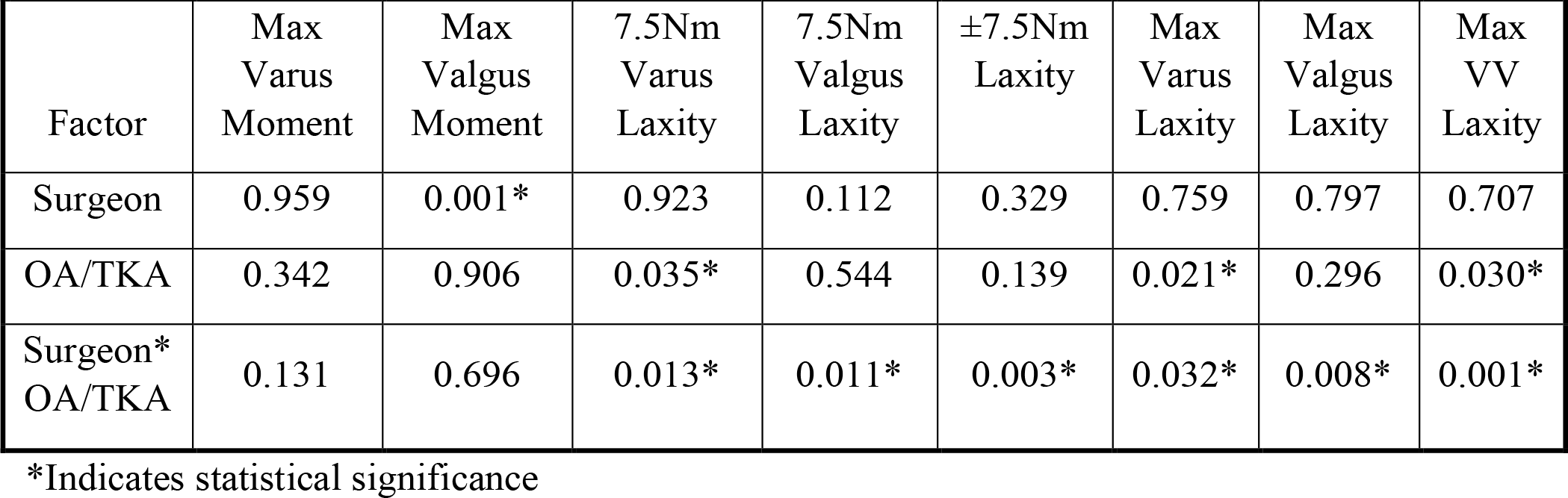
Laxity Analysis of Variance By Surgeon and OA vs. TKA Knee. (P Values)

The manner in which TKA laxity was achieved differed by surgeon, as indicated by the significant surgeon x OA/TKA interaction effect (Tables 1-3, Figure 4). For example, under the ±7.5 N·m varus-valgus applied load, Surgeon 1 “tightened” knees by as much as 1.9° and “loosened” knees by as much as 5.8°, while Surgeon 2 “tightened” knees by as much as 6.7° and “loosened” them by as much as 5.4°(Figure 4). Under the maximum applied load, Surgeon 1 “tightened” knees by as much as 4.0° and “loosened” knees by as much as 9.5°, while Surgeon 2 “tightened” knees by as much as 2.8° and “loosened” knees by as much as 5.0°

**Figure 4:**
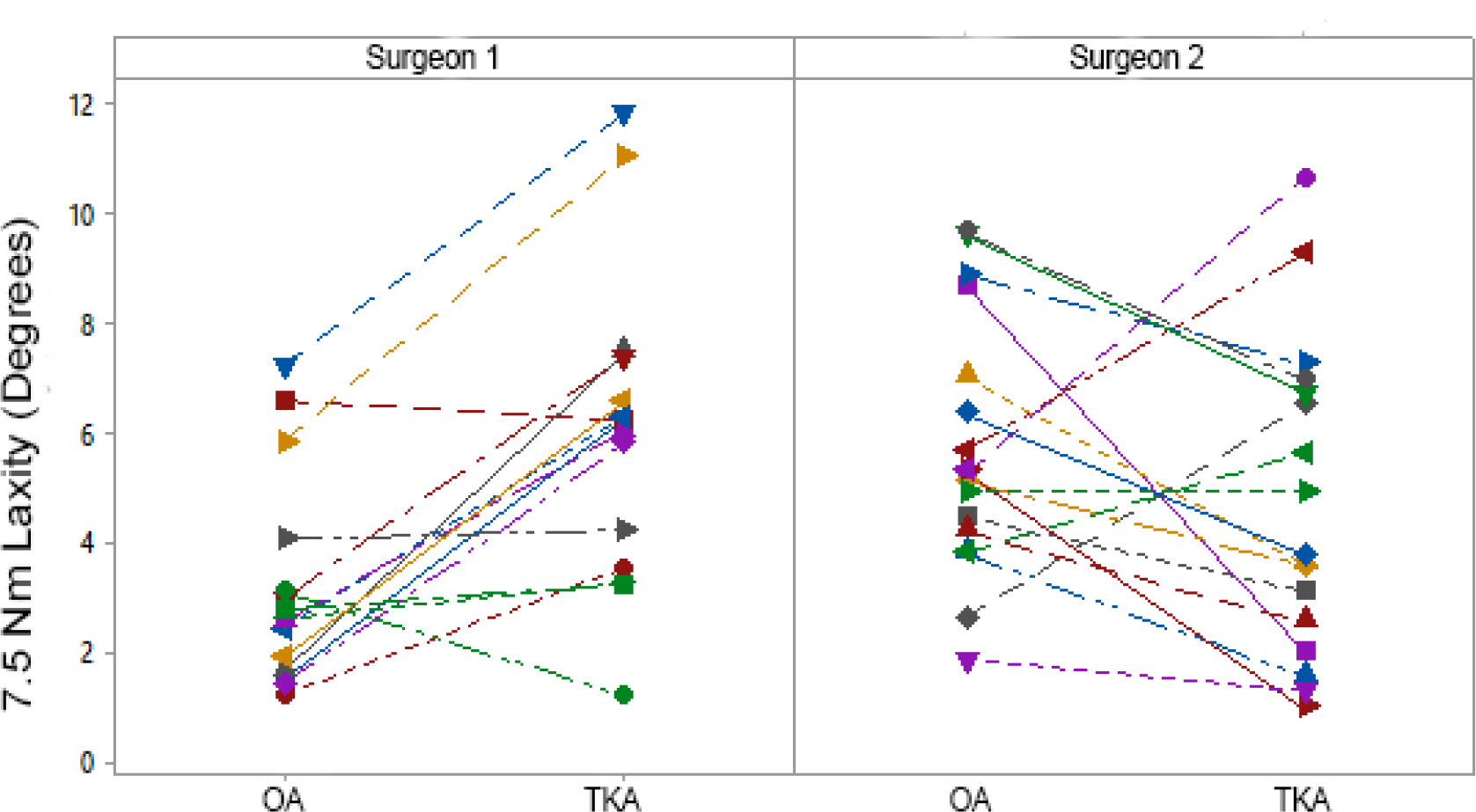
Differences in ± 7.5 N·m laxity in the OA and TKA knees for the two surgeons. Each icon represents an individual patient, with the lines connecting the OA and TKA measurements. Surgeon 1 generally loosened knees and left knees with more laxity on average than Surgeon 2.

### Objectively determine whether TKA resulted in a balanced knee

The knees recorded in the operating room were balanced post-operatively on average, with no differences seen between varus and valgus laxity at 7.5 N·m (0.16° ± 1.88° varus bias) (p = 0.629) or at the maximum applied moment (0.03° ± 3.05° varus bias) (p = 0.941). There were no differences in balance between surgeons at either 7.5 N·m (Table 2) or the maximum applied loads (Table 3)

**Table 2:** Comparison of Laxity and Balance Between the 2 Surgeons in OA and TKA Knees Under 7.5 N·m Applied Loads. P-values Indicate Significant Differences Between the Cohorts of Each Surgeon and between the OA and TKA conditions for each surgeon

| Measurement | Surgeon 1<br>Mean ± STD | Surgeon 1<br>Range | Surgeon 2<br>Mean ± STD | Surgeon 2<br>Range | Surgeon 1 vs 2<br>p-value |
| --- | --- | --- | --- | --- | --- |
| OA 7.5 N·m<br>Varus laxity | 1.7°±1.1° | 0.4°- 4.0° | 2.5°±1.1° | 0.7°- 5.4° | p=0.052 |
| TKA 7.5 N·m<br>Varus laxity | 3.2°±1.7° | 0.6°- 7.3° | 2.5°±1.6° | 0.6°- 6.0° | p=0.214 |
| p-value<br>OA vs. TKA<br>Varus laxity | p = 0.006 * |  | p=0.612 |  |  |
| OA 7.5 N·m<br>Valgus laxity | 1.5°±1.0° | 0.7°- 4.1° | 3.2°±1.5° | 1.1°- 6.3° | p < 0.001 * |
| TKA 7.5 N·m<br>Valgus laxity | 2.8°±2.0° | 0.5°- 9.2° | 2.4°±1.6° | 0.0°- 5.5° | p=0.548 |
| p-value<br>OA vs. TKA<br>Valgus laxity | p = 0.010 * |  | p=0.124 |  |  |
| OA ±7.5 N·m<br>Total laxity | 3.2°±1.9° | 1.2°- 7.2° | 5.7 ± 2.3° | 1.9°- 9.7° | p = 0.002 * |
| TKA ±7.5 N·m<br>Total laxity | 6.0°±2.8° | 1.2°- 11.8° | 4.7°±2.8°) | 1.0°- 10.6°) | p=0.208 |
| p-value<br>OA vs. TKA<br>Total laxity | P < 0.001 |  | p=0.209 |  |  |
| OA ±7.5 N·m<br>Laxity Balance | 0.20° ± 0.84°<br>varus bias | 1.5° of more<br>valgus motion<br>to 2.2° of<br>more varus<br>motion | 0.67° ± 1.45°<br>valgus bias | 3.2° of more<br>valgus motion<br>to 2.2° of<br>more varus<br>motion | p = 0.041 * |
| TKA ±7.5 N·m<br>Laxity Balance | 0.42° ± 2.49°<br>varus bias | 6.6° of more<br>valgus motion<br>to 3.8° of<br>more varus<br>motion | 0.06° ± 1.20°<br>valgus bias | 2.0° of more<br>valgus motion<br>to 2.0° of<br>more varus<br>motion | p=0.508 |
| p-value<br>OA vs. TKA<br>Laxity Balance | p=0.738 |  | p=0.226 |  |  |

**Table 3:**
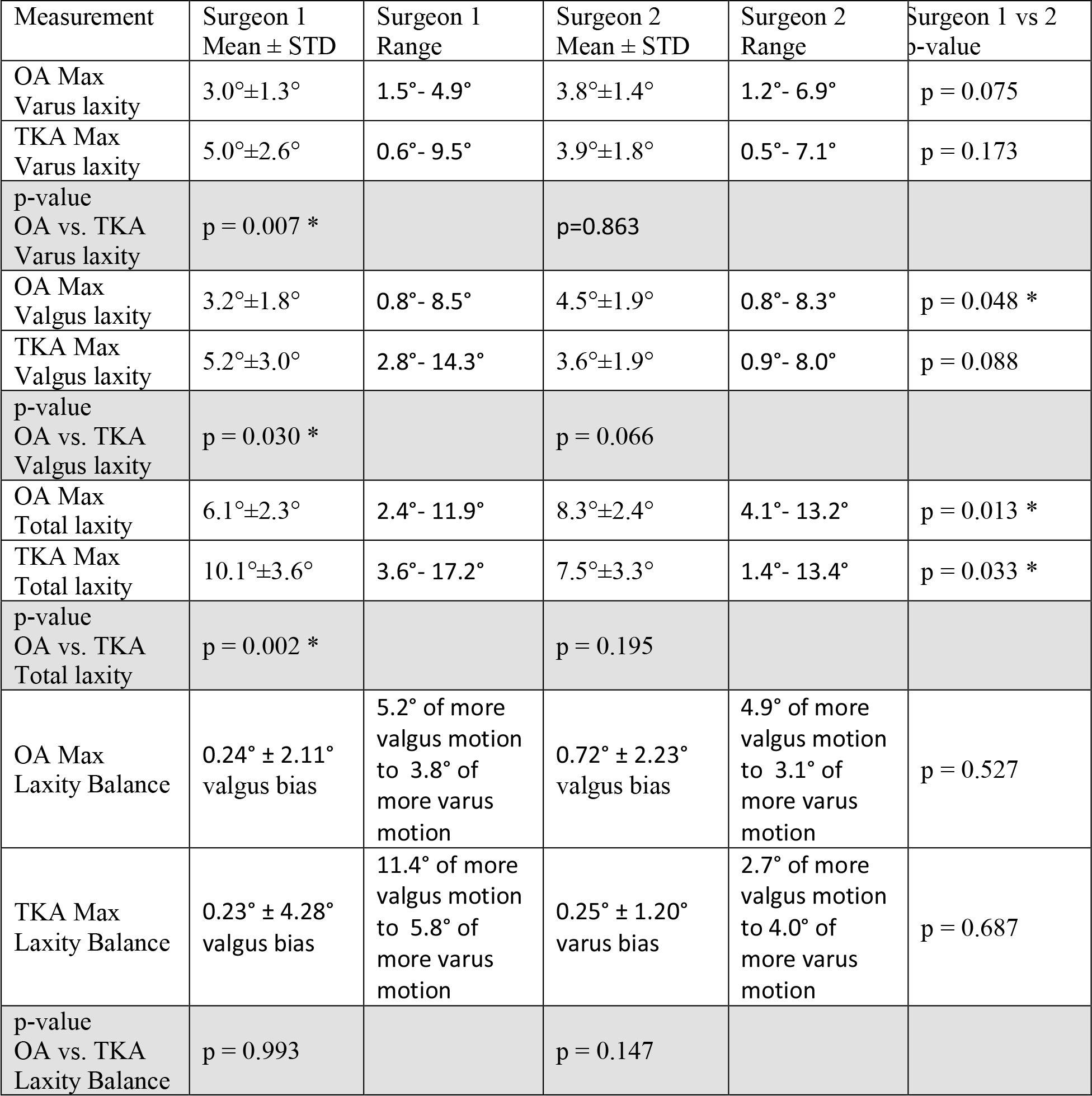
Comparison of Laxity and Balance Between the 2 Surgeons in OA and TKA Knees Under Maximum Applied Loads. P-values Indicate Significant Differences Between the Cohorts of Each Surgeon and between the OA and TKA conditions for each surgeon

### Compare measurements of TKA laxity against values from cadaveric knees

Several significant differences were seen between the intraoperative measurements and the “control” data from the cadaveric specimens (Table 4, Figure 5). Under a ±7.5 Nm load, varus laxity, valgus laxity, and varus-valgus laxity in both the OA and TKA knees were different from the cadaveric specimens. However, at this load, there was no difference in balance between either the OA knees (0.28° ± 1.27° valgus bias) or TKA knees (0.16° ± 1.88° varus bias) and the cadaveric knees (0.25° ± 0.66° varus bias). Under the maximum applied load, we observed many of the same differences in the laxity parameters between the OA or TKA knees and the cadaveric specimens. Only two comparisons were not statistically different: the maximum varus between the OA and cadaveric knees and the balance between the TKA and cadaveric knees.

**Table 4:** Comparison of Laxity and Balance in OA and TKA Knees to Native Cadaveric Knees

| Measurement | OA<br>Mean ± STD | p-value<br>OA-Cadaver | Cadaver<br>Mean ± STD | p-value<br>TKA-Cadaver | TKA<br>Mean ± STD |
| --- | --- | --- | --- | --- | --- |
| 7.5 N·m<br>Varus laxity | 2.13° ± 1.19° | p = 0.001 * | 1.16° ± 1.08° | p < 0.001 * | 2.81° ± 1.69° |
| 7.5 N·m<br>Valgus laxity | 2.41° ± 1.55° | p < 0.001 * | 0.92° ± 0.58° | p < 0.001 * | 2.57° ± 1.82° |
| ±7.5 N·m<br>Total laxity | 4.55° ± 2.45° | p < 0.001 * | 2.08° ± 1.60° | p < 0.001 * | 5.33° ± 2.86° |
| ±7.5 N·m<br>Laxity<br>Balance | 0.28° ± 1.27°<br>Valgus bias | p = 0.037 | 0.25° ± 0.66°<br>Varus bias | p = 0.804 | 0.16° ± 1.88°<br>Varus bias |
| Maximum<br>Varus laxity | 3.42° ± 1.37° | p = 0.086 | 2.83° ± 1.53° | p = 0.001 * | 4.34° ± 2.22° |
| Maximum<br>Valgus laxity | 3.92° ± 1.93° | p < 0.001 * | 2.25° ± 1.20° | p < 0.001 * | 4.31° ± 2.52° |
| Maximum<br>Total laxity | 7.34° ± 2.56° | p < 0.001 * | 5.08° ± 2.60° | p < 0.001 * | 8.65° ± 3.65° |
| Maximum<br>Laxity<br>Balance | 0.51° ± 2.16°<br>Valgus bias | p = 0.008 * | 0.59° ± 0.90°<br>Varus bias | p = 0.317 | 0.04° ± 3.05°<br>Varus bias |

**Figure 5:**
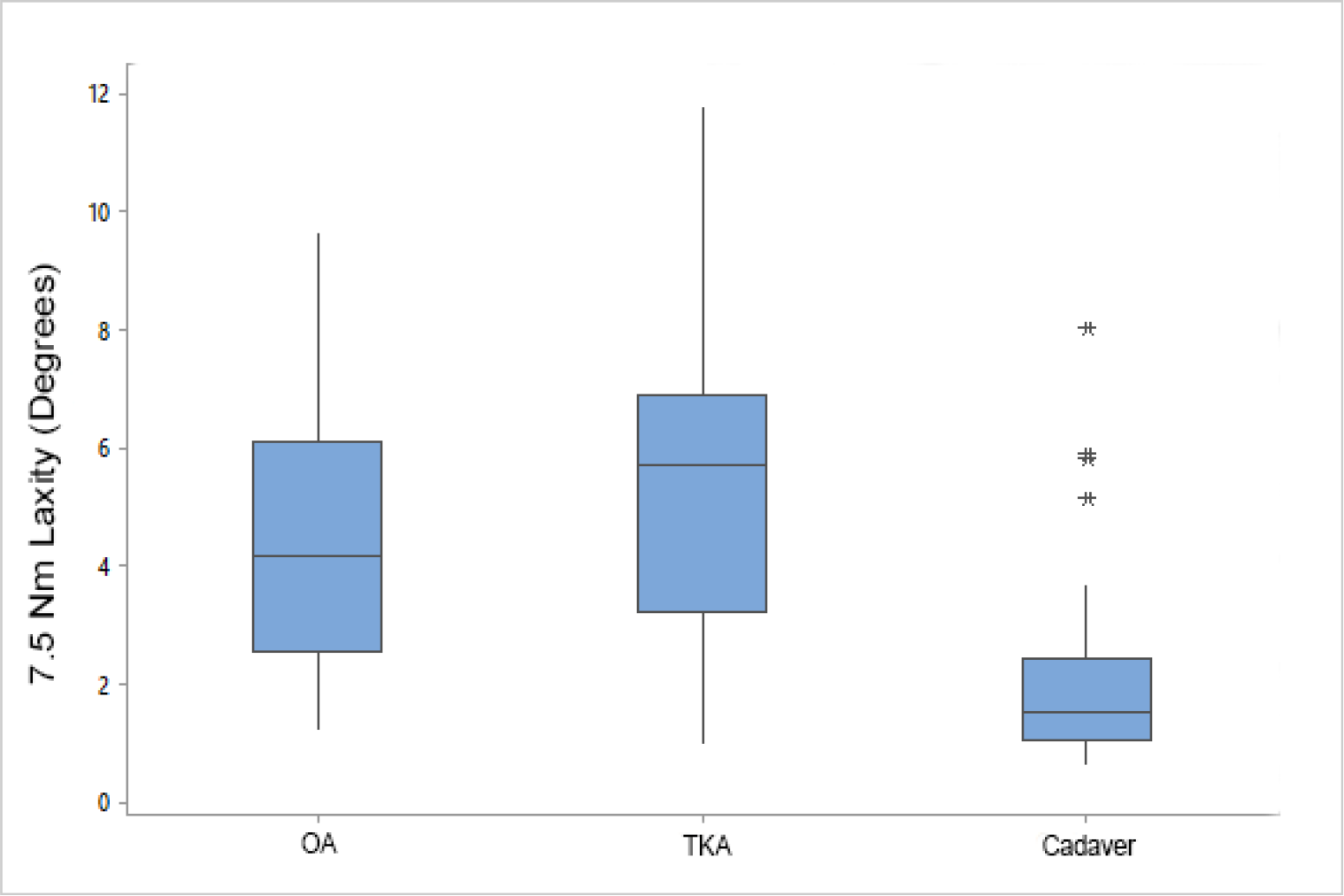
Differences in ±7.5 N·m laxity in the OA, TKA, Cadaver knees. Both OA and TKA kness were more lax than the “normal” cadaveric specimens.

## Discussion

We believe this study represents the first objective characterization of intra-operative knee laxity and balance in response to a measured applied load before and after TKA, whereas other approaches can only be used after some bone cuts have been performed. Further, data suggest that even balanced knees can have different amounts of knee laxity.

Our results generally agree with and build upon previous research into knee laxity. We previously demonstrated that TKA results in greater knee laxity in cadavers with PS implants [29] and in patients the operating room in response to an unmeasured load[39]. Our data from the cadavers in the present study are similar to other laxity data in cadavers[25] and human subjects[40] and support the conclusion that knees with osteoarthritis are more lax than healthy controls[41]. However, our finding of a biomechanically balanced knee contrasts other work whic found unbalanced (i.e., asymmetric) knees in full extension measured with stress radiographs [9]. This study raises an interesting question as to how post-operative laxity is established with TKA. Even though, on average, the varus-valgus laxity and balance for the TKA knees were similar between the 2 surgeons and was looser than the OA knees, Surgeon 1 tended to “loosen” a cohort of patients with “tighter” knees, while Surgeon 2 did not appreciably alter varus-valgus laxity. Both surgeons loosened and tightened knees within their respective cohorts, even though they had similar approaches to the operation. Thus, it remains unknown whether our results are due to the desire of both surgeons to leave the knees in a similar state post-operatively or whether our results would be changed if the surgeons were presented with a different cohort of patients and they continued their general approach of either loosening or not altering laxity.

Similar to how navigation systems have been used to document the variability with component alignment [42], our data suggest that large variability exists in what is considered acceptable laxity and balance of the TKA knee when it is assessed by only manual manipulation. The varus/valgus loads applied to some patients were almost 10 times the loads that were applied to others. With post-implant ±7.5 N·m varus-valgus laxity varying from 1.0° to 11.8°, maximum varus-valgus laxity varying from 1.4° to 17.2°, and magnitudes of balance varying by over 11°, it is clear that even though all patients received the same implant, the overall state of their knees were greatly different from each other. It is important to question whether such variability is clinically meaningful. Previous studies have found statistically significant differences in varus-valgus laxity of 1.5° when measured at 20° of knee flexion between OA patients and healthy older control subjects[41], have used a threshold of 4° as the difference between “tight” and “slack” TKA knees when measured at 30° of knee flexion[43], and have a threshold of 6° to deduct points from the Knee Society Score with the knee is in full extension after TKA [44]. Given this variability in the TKA knees, the initial variability of the pre-operative (OA) knees seen by the surgeons, as well as how different surgeons affected knee laxity, personalized approaches to performing a TKA that involve objective measurements of surgical technique may be warranted to ensure consistent post-operative outcomes.

This study is not without limitations. We only tested 34 knees with similar pre-operative deformities, all of whom underwent TKA by the same two surgeons, and using the same implant tested in full extension. Different surgeons using a different surgical approach (such as the so-called “kinematic alignment” approach[45]), different implants (different manufacturer, cruciate retaining designs or medially stabilized implants [46, 47]), a different cohort of patients (e.g., valgus deformity), would demonstrate different results. Additionally, knees in this initial cohort were only tested in full-extension due the learning curve associated with the time recording these measurements; testing at different flexion angles would yield different results. Despite these limitations, we believe that this initial study demonstates a more objective and comprehensive characterization of intra-operative joint laxity than has historically been possible.

In conclusion, we have demonstrated that while knees following TKA had equal amounts of varus-valgus motion under load (i.e., was “balanced”), “normal” joint laxity is not achieved and the OA (pre-operative) laxity conditions of the joint may not be retained. It is unknown if TKA should attempt to restore the pre-operative condition of the joint, should establish “biomechanical balance,” should establish the “normal” non-OA knee laxity conditions, or leave the knee in another condition. Future work should relate objective definitions of the biomechanics of the knee to clinical outcomes in order to improve patient satisfaction and post-operative functionality.

## Data Availability

data is not shared publicly

